# Genomic heterogeneity and clinical characterization of SARS-CoV-2 in Oregon

**DOI:** 10.1101/2020.07.30.20160069

**Authors:** Alexa K Dowdell, Kevin Matlock, Fred L. Robinson, Roshanthi Weerasinghe, Rogan Rattray, Marina Pukay, Melvin Lathara, Anastacia Harlan, Thomas R. Ward, Mary Campbell, Walter Urba, Ganapati Srinivasa, Carlo B. Bifulco, Brian D. Piening

## Abstract

The first reported case of COVID-19 in the State of Oregon occurred in late February 2020, with subsequent outbreaks occurring in the populous Portland metro area but also with significant outbreaks in less-populous and rural areas. Here we report viral sequences from 188 patients across the hospitals and associated clinics in the Providence Health System in the State of Oregon dating back to the early days of the outbreak. We show a significant shift in dominant clade lineages over time in Oregon, with the rapid emergence and dominance of Spike D614G-positive variants. We also highlight significant diversity in SARS-CoV-2 sequences in Oregon, including a large number of rare mutations, indicative that these genomes could be utilized for outbreak tracing. Lastly, we show that SARS-CoV-2 genomic information may offer additional utility in combination with clinical covariates in the prediction of acute disease phenotypes.

## INTRODUCTION

Since late 2019 the COVID-19 pandemic has rapidly spread across the globe, with over 12 million reported cases worldwide as of July 2020. In a number of the hardest-hit regions this has resulted in overwhelmed intensive care units, ventilator shortages and significant morbidity and mortality. While a variety of novel therapeutic strategies have shown different degrees of efficacy, a vaccine is not currently available, making infection avoidance measures such as social distancing and universal masking key strategies in reducing infection and death rates.

In the United States there is significant heterogeneity in SARS-CoV-2 spread at the state and county level due to differences in population density, coordinated preventive measures, ability to perform widespread testing and coordinated outbreak response and tracing. Testing for SARS-CoV-2 infection has almost exclusively been performed via real-time reverse transcriptase polymerase chain reaction (RT-PCR) due to high multiplexability of these assays, rapid turnaround time to result, relatively low cost and availability of RT-PCR platforms in clinical laboratory settings. However, most of these assays provide a qualitative yes/no result on whether SARS-CoV-2 is present or not, and do not provide additional information about the genomic sequence of the particular SARS-CoV-2 strain.

Genomic sequencing is rapidly emerging as an orthogonal strategy to RT-PCR for COVID-19 outbreak monitoring as the sequence specificity uncovered in individual SARS-CoV-2 isolates has shown significant utility for the epidemiological investigation of outbreak origins as well as the early identification of possible functional changes to the virus that may affect transmission rates or associated clinical outcomes. For example, Bedford et al. sequenced hundreds of genomes in Washington State (WA) to uncover early cryptic spread of SARS-CoV-2 likely due to a single progenitor event (Bedford et al., 2020). In contrast, work from Chiu and colleagues showed genomic evidence for up to eight early and diverse introduction events in Northern California (CA) (Deng et al., 2020). More recently, work from multiple groups have utilized SARS-CoV-2 sequence data to identify a more infectious circulating strain of SARS-CoV-2 hallmarked by a characteristic D614G alteration in the spike protein (Zhang et al., 2020).

While genomic sequencing has been utilized to effectively trace origins of SARS-CoV-2 in WA and CA, in neighboring Oregon (OR) the picture is less clear, partly due to a paucity of available genomic sequences as well as associated clinical information. We performed large-scale sequencing of SARS-CoV-2 genomes isolated from 188 patients across the largest healthcare system in Oregon. In this context, we show significant diversity across the scope of SARS-CoV-2 genomes in OR as well as potential links to clinical phenotypes in this cohort.

## RESULTS

### Cohort Overview

In order to assess the heterogeneity of SARS-CoV-2 genomes across OR, a total of 204 nasopharyngeal swab patient specimens (representing 188 unique patients) were sequenced from diverse clinical sites across OR. A total of 179 of these resulted in complete SARS-CoV-2 genome coverage (average-per-base coverage of 60x or greater) and were included for further study. For a subset of patients, virus from multiple longitudinal swab specimens were sequenced in order to determine whether any novel mutations emerge within an individual over the course of COVID-19. The demographics of the tested patients are reported in **Table 1**. Specifically, twelve of the patients died during the course of COVID-19, seven required intubation and 54 were admitted for acute or intensive care.

**Table 1.**
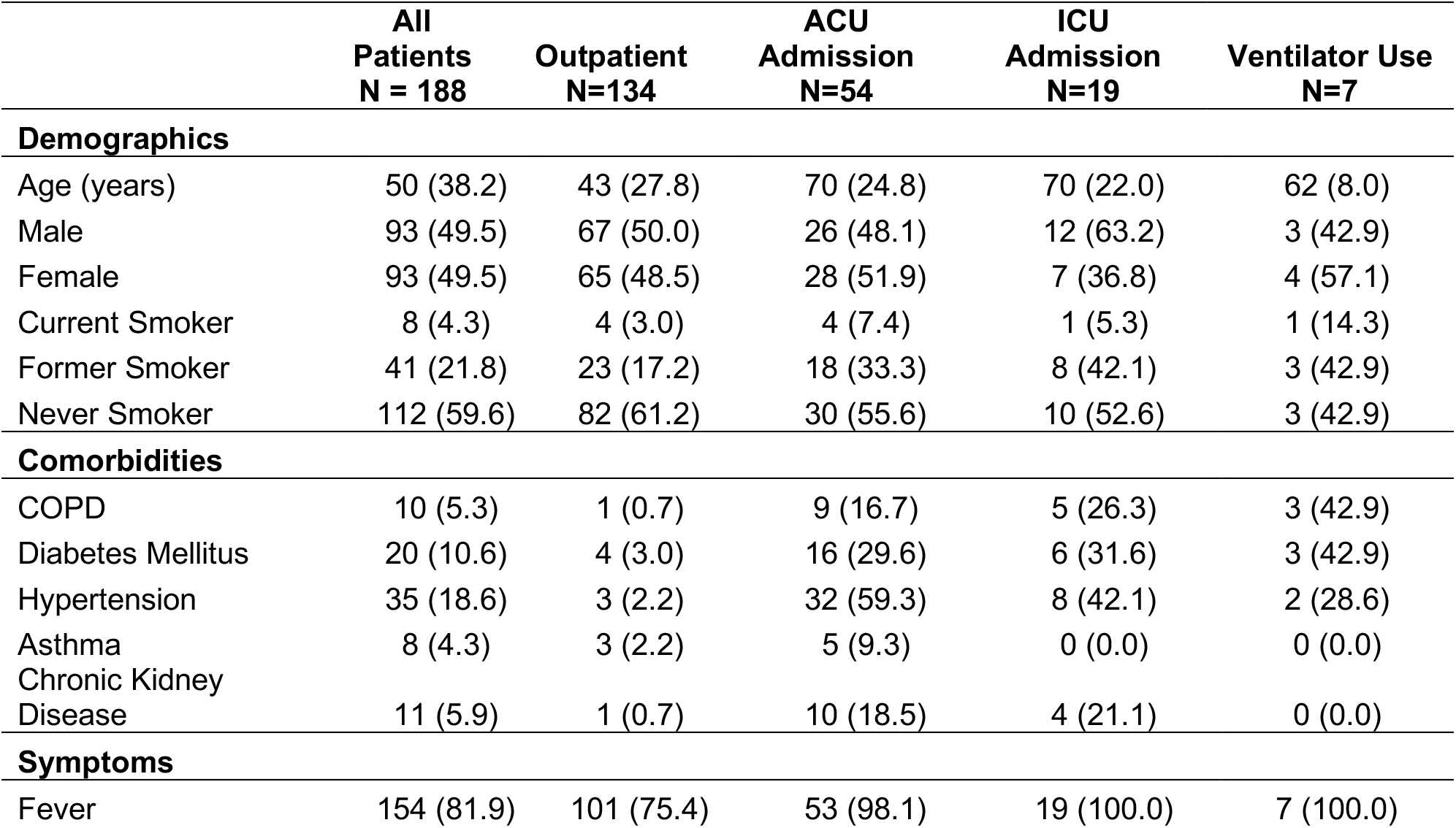
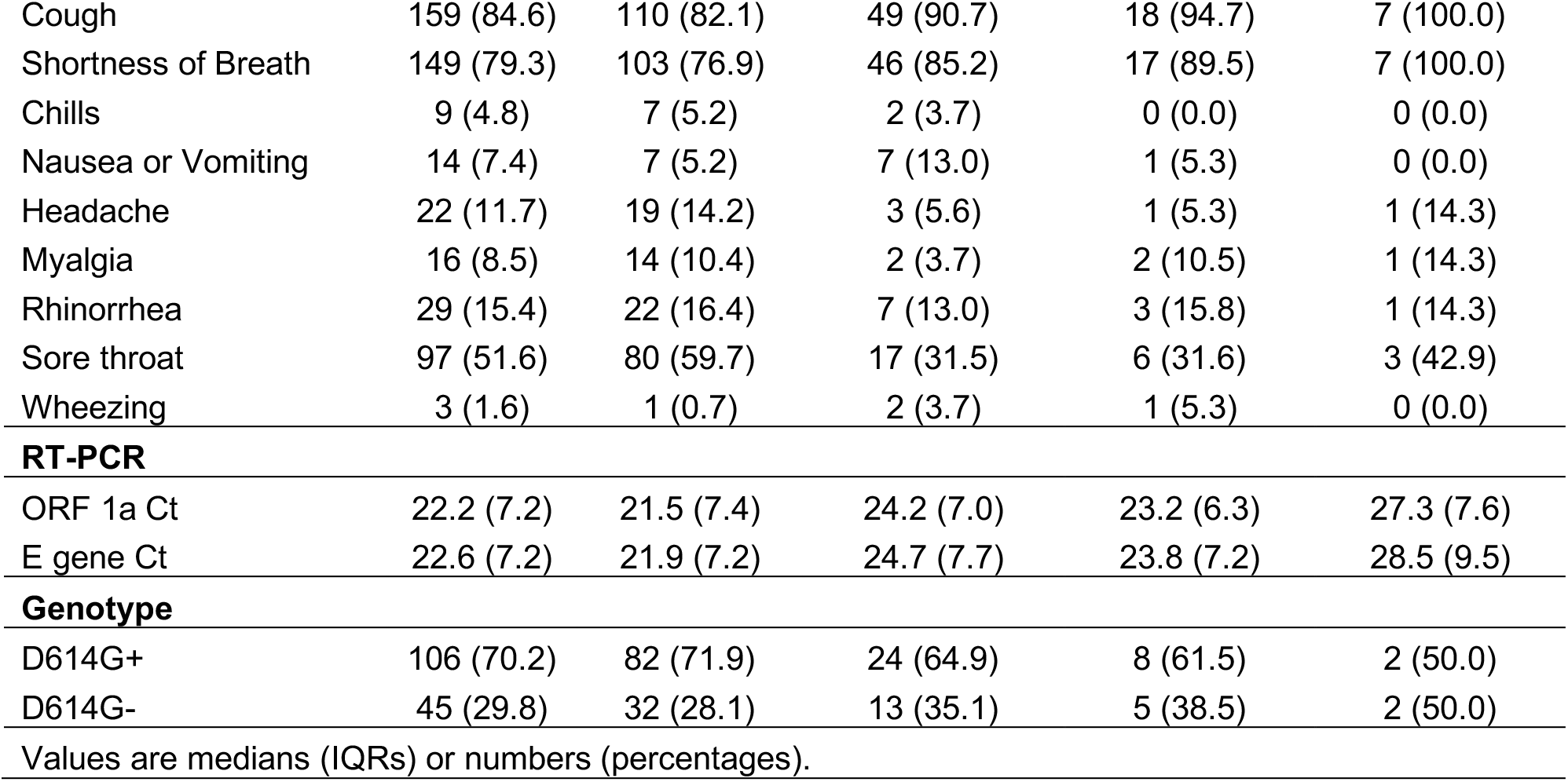
Demographic, Comorbidities, Symptoms, RT-PCR Thresholds, and Viral Genotypes of Patient Dataset stratified across Outcomes.

|  | All<br>Patients<br>N = 188 | Outpatient<br>N=134 | ACU<br>Admission<br>N=54 | ICU<br>Admission<br>N=19 | Ventilator Use<br>N=7 |
| --- | --- | --- | --- | --- | --- |
| **Demographics** |  |  |  |  |  |
| Age (years) | 50 (38.2) | 43 (27.8) | 70 (24.8) | 70 (22.0) | 62 (8.0) |
| Male | 93 (49.5) | 67 (50.0) | 26 (48.1) | 12 (63.2) | 3 (42.9) |
| Female | 93 (49.5) | 65 (48.5) | 28 (51.9) | 7 (36.8) | 4 (57.1) |
| Current Smoker | 8 (4.3) | 4 (3.0) | 4 (7.4) | 1 (5.3) | 1 (14.3) |
| Former Smoker | 41 (21.8) | 23 (17.2) | 18 (33.3) | 8 (42.1) | 3 (42.9) |
| Never Smoker | 112 (59.6) | 82 (61.2) | 30 (55.6) | 10 (52.6) | 3 (42.9) |
| **Comorbidities** |  |  |  |  |  |
| COPD | 10 (5.3) | 1 (0.7) | 9 (16.7) | 5 (26.3) | 3 (42.9) |
| Diabetes Mellitus | 20 (10.6) | 4 (3.0) | 16 (29.6) | 6 (31.6) | 3 (42.9) |
| Hypertension | 35 (18.6) | 3 (2.2) | 32 (59.3) | 8 (42.1) | 2 (28.6) |
| Asthma | 8 (4.3) | 3 (2.2) | 5 (9.3) | 0 (0.0) | 0 (0.0) |
| Chronic Kidney Disease | 11 (5.9) | 1 (0.7) | 10 (18.5) | 4 (21.1) | 0 (0.0) |
| **Symptoms** |  |  |  |  |  |
| Fever | 154 (81.9) | 101 (75.4) | 53 (98.1) | 19 (100.0) | 7 (100.0) |
| Cough | 159 (84.6) | 110 (82.1) | 49 (90.7) | 18 (94.7) | 7 (100.0) |
| Shortness of Breath | 149 (79.3) | 103 (76.9) | 46 (85.2) | 17 (89.5) | 7 (100.0) |
| Chills | 9 (4.8) | 7 (5.2) | 2 (3.7) | 0 (0.0) | 0 (0.0) |
| Nausea or Vomiting | 14 (7.4) | 7 (5.2) | 7 (13.0) | 1 (5.3) | 0 (0.0) |
| Headache | 22 (11.7) | 19 (14.2) | 3 (5.6) | 1 (5.3) | 1 (14.3) |
| Myalgia | 16 (8.5) | 14 (10.4) | 2 (3.7) | 2 (10.5) | 1 (14.3) |
| Rhinorrhea | 29 (15.4) | 22 (16.4) | 7 (13.0) | 3 (15.8) | 1 (14.3) |
| Sore throat | 97 (51.6) | 80 (59.7) | 17 (31.5) | 6 (31.6) | 3 (42.9) |
| Wheezing | 3 (1.6) | 1 (0.7) | 2 (3.7) | 1 (5.3) | 0 (0.0) |
| **RT-PCR** |  |  |  |  |  |
| ORF 1a Ct | 22.2 (7.2) | 21.5 (7.4) | 24.2 (7.0) | 23.2 (6.3) | 27.3 (7.6) |
| E gene Ct | 22.6 (7.2) | 21.9 (7.2) | 24.7 (7.7) | 23.8 (7.2) | 28.5 (9.5) |
| **Genotype** |  |  |  |  |  |
| D614G+ | 106 (70.2) | 82 (71.9) | 24 (64.9) | 8 (61.5) | 2 (50.0) |
| D614G- | 45 (29.8) | 32 (28.1) | 13 (35.1) | 5 (38.5) | 2 (50.0) |

Values are medians (IQRs) or numbers (percentages).

### Genomic Diversity of the OR Cohort

Sequence results from the 179 OR specimens revealed a striking diversity of SARS-CoV-2 genomes across Oregon (**Figure 1A**). Virtually all major worldwide GISAID clades (Shu and McCauley, 2017) are represented in OR genomes, with the largest number in GH and G as well as sizable clusters in S and V-clades. Of note the former major clades have highest representation in Europe, but were also observed as the dominant clades in the spread to South America, Africa and Canada. The S-clade exhibits highest representation in Asia and V-clade represents an early introduction of the virus into Italy and Europe from China (Giovanetti et al., 2020). Genomic variation in these isolates spanned the entirety of the SARS-CoV-2 genome with notable hotspots in *ORF1A, ORF3A* and *Spike* (**Figure 1B**), and variant classes were predominantly missense or synonymous (**Figure 1C**). A cohort of isolates mapped to S-clade with high similarity to the original WA isolate (one of the earliest introduction events in the United States (Holshue et al., 2020)) as well as other early WA-specific sequences (Bedford et al., 2020). Given the extent of frequent travel between OR and WA, it is perhaps unsurprising that a large number of early isolates bear similarity to WA SARS-CoV-2 sequences. Overall, we observed a shift in clade representation over time, with the early cases dominated primarily by L-, S- and V-clades and later cases predominated by the G-clades (**Figure 2A**). Of particular note for outbreak tracing, we note that even within clades there is a striking amount of sequence diversity, with 55% of SARS-CoV-2 genomes containing at least one nucleotide variant that was completely unique among the cohort, and an additional 19% that shared an identical SARS-CoV-2 genome with only one other patient (**Figure 2B**). Only twelve percent of the genomes were common among four or more patients in the cohort.

**Figure 1.**
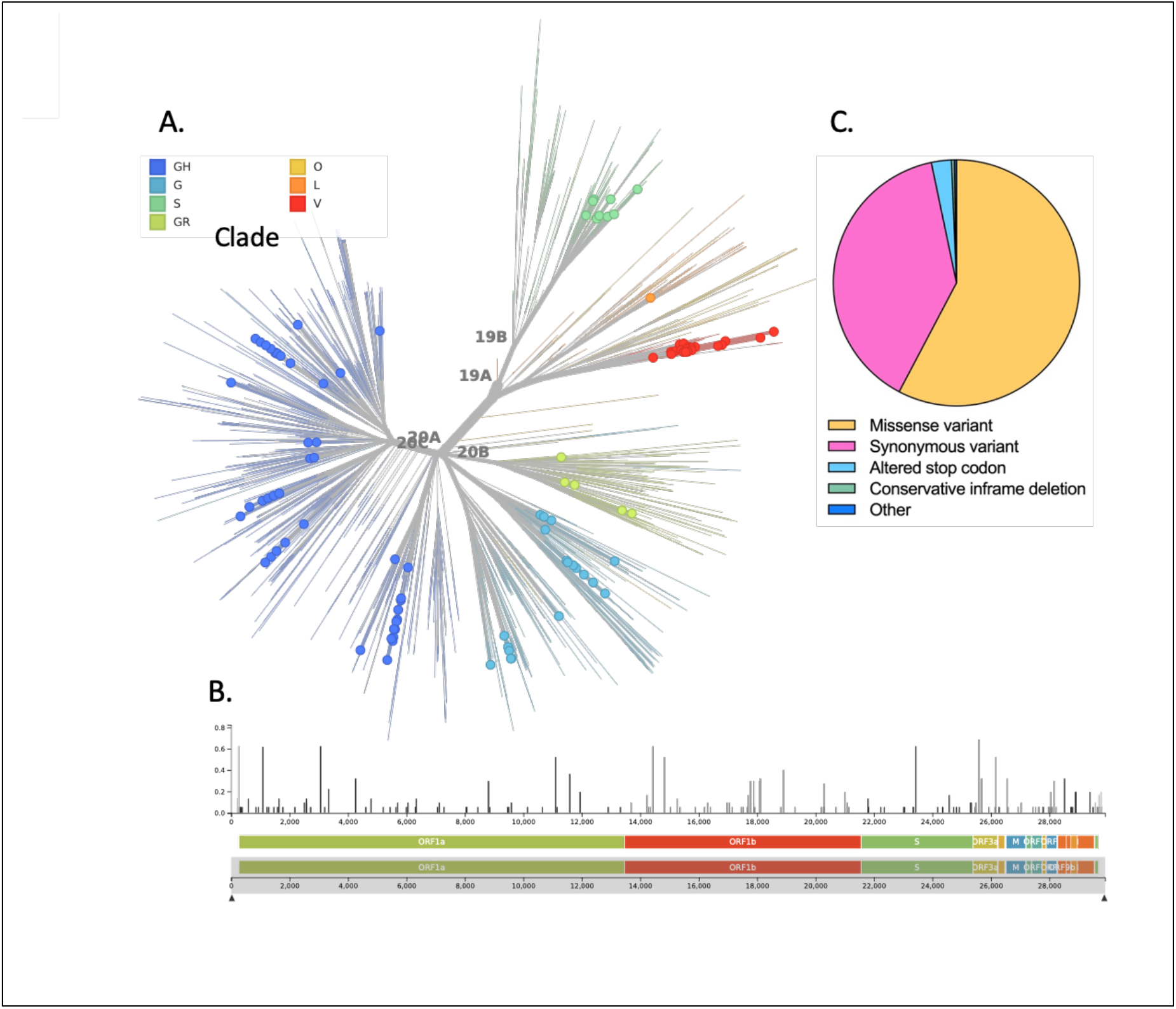
Genomic diversity of SARS-CoV-2 in Oregon. **A)** Unrooted phylogenetic tree of major SARS-CoV-2 clades with representative worldwide genomes subsampled from GISAID. OR isolates sequenced in this study are represented by circles. **B)** Genomic location and histogram of SARS-CoV-2 mutations across the OR isolates. **C)** Distribution of variant effects across the cohort.

**Figure 2.**
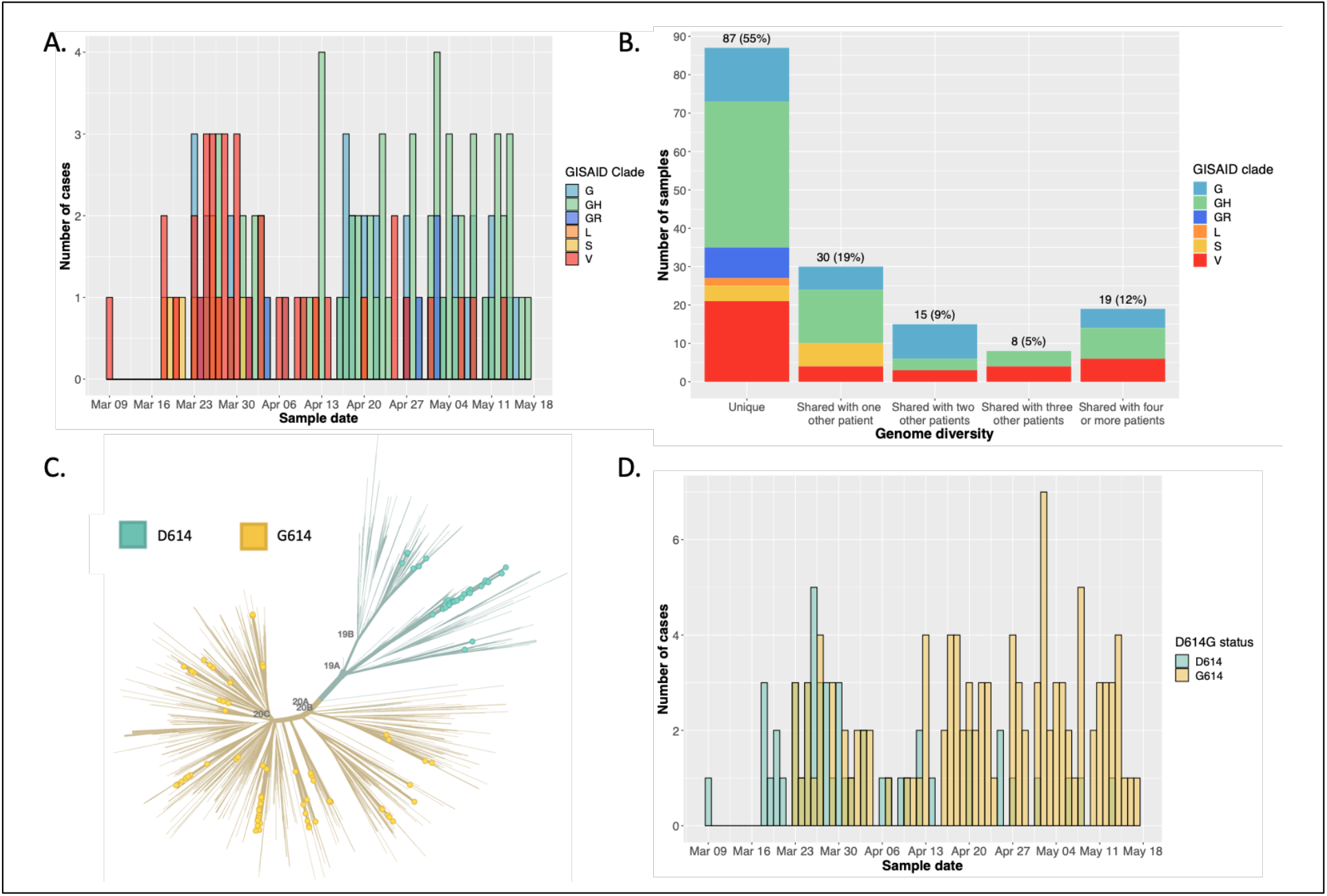
SARS-CoV-2 genotype diversity over time in OR. **A)** Histogram of clade representation by date for the Oregon SARS-CoV-2 genomes. **B)** Distribution of SARS-CoV-2 genomes that are unique to a single patient, shared among only two patients, shared among three and four or more patients. **C)** Phylogenetic tree showing distribution of D614 or G614 Spike genotype. **D)** Histogram of D614G status of Oregon SARS-CoV-2 genomes over time.

The identification of known functional variants in SARS-CoV-2 is of particular importance, especially given recent data showing increased infectivity and rapid spread of the D614G variant across the United States and worldwide. Our data show that D614G is already the dominant genotype in Oregon, with 69% of sequenced genomes coding the Glycine substitution (**Figure 2C**). Notably, in Oregon this variant emerged over time as it was present at a significantly higher proportion in the second half of the tested period (Mid-April to May) (p<1⨯10^−8^) (**Figure 2D**). We also tracked prevalence of D614G at specific subregions in OR based on clinic of origin (**Figure 3**). Notably, D614G prevalence varied dramatically by region, with sites near Hillsboro and Newberg exhibiting 50% or below prevalence of D614G and sites east of the Willamette river exhibiting a very high prevalence of D614G.

**Figure 3.**
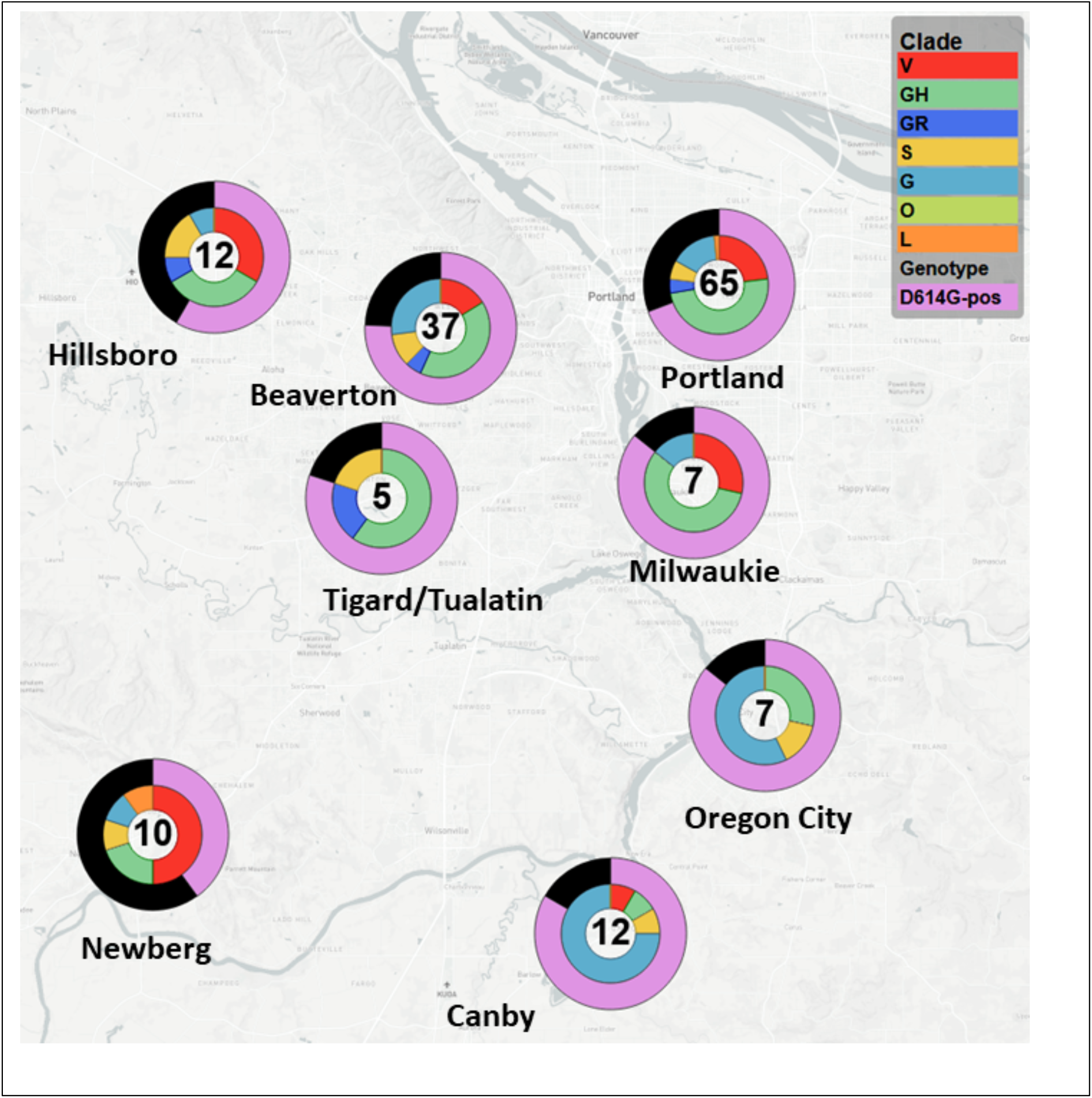
SARS-CoV-2 genotype diversity over time in OR. Distribution of SARS-CoV-2 clade (inner pie distribution, see key) as well as D614 or G614 Spike genotype (outer pie distribution) at individual sample isolation sites across the Portland metro region. Number of samples is indicated in the center of each circle.

We also identified two rare mutations in the receptor binding motif (RBM) of the SARS-CoV-2 Spike protein (A475V and P463S). This segment of Spike mediates interaction with ACE2 (Lan et al., 2020; Shang et al., 2020; Wang et al., 2020; Yan et al., 2020). Structural studies indicate that Ala-475 of Spike engages in a hydrogen bond with Ser-19 of ACE2 (Lan et al., 2020; Shang et al., 2020; Wang et al., 2020). A second rare mutation in the RBM of Spike, P463S, is located in a highly-conserved residue that does not directly contact ACE2, but may be involved in maintaining the overall conformation of the RBM (Lan et al., 2020; Shang et al., 2020; Wang et al., 2020; Yan et al., 2020). Given the importance of Spike-ACE2 binding in viral cell entry, follow-up studies on the impact of these variants may be warranted. Mutations with the potential to alter the function of the RBM may be of relevance for infectivity, and might also impact the function of neutralizing antibodies recognizing Spike.

### Associations Between SARS-CoV-2 Genotype and Patient Clinical Characteristics

Next, we sought to determine whether sequence variants in the isolated SARS-CoV-2 genomes were associated with differential clinical manifestation of COVID-19 in the patients. We looked at associations of SARS-CoV-2 variants with level of clinical care required; this included entry into an Acute Care Unit (ACU), Intensive Care Unit (ICU) or placement on a ventilator (**Figure 4**). This was done in comparison with other demographic and prior comorbidities as acute care predictors. A list of these characteristics along with a statistical summary across the different clinical categories is provided in **Table 1**. We note that despite the high prevalence of SARS-CoV-2 D614G in our study cohort, only one of twelve deceased patients had a D614G-positive infection (p<0.001, Fisher’s exact test). Despite this, D614G genotype did not well differentiate between overall admission status in the cohort. In contrast, as expected we see a strong differentiation between the ages of Outpatient vs ACU admitted patients (*p* =3.72*e*-10Mann-Whitney U Test), along with current or former smoking status (*p* = 7.75*e*-3 G-Test). Comorbidities were the strongest indicator of hospital admission, with all but asthma having a *p* ≤ 1.79*e*-3 (G-Test). The p-value for asthma was *p* = 9.63*e*-2 however we note the small sample size of this population. Finally, SARS-CoV-2 RT-PCR Ct values are overall higher in the admitted patient group (*p* = 2.97*e*-4 Mann-Whitney U Test), suggestive of an inverse relationship between nasal viral titer and disease severity. Applying unsupervised Uniform Manifold Approximation and Projection (UMAP) using the clinical and genomic variables revealed several distinct clusters, however none of these clearly separated outpatient vs admitted patients (**Figure 5**).

**Figure 4.**
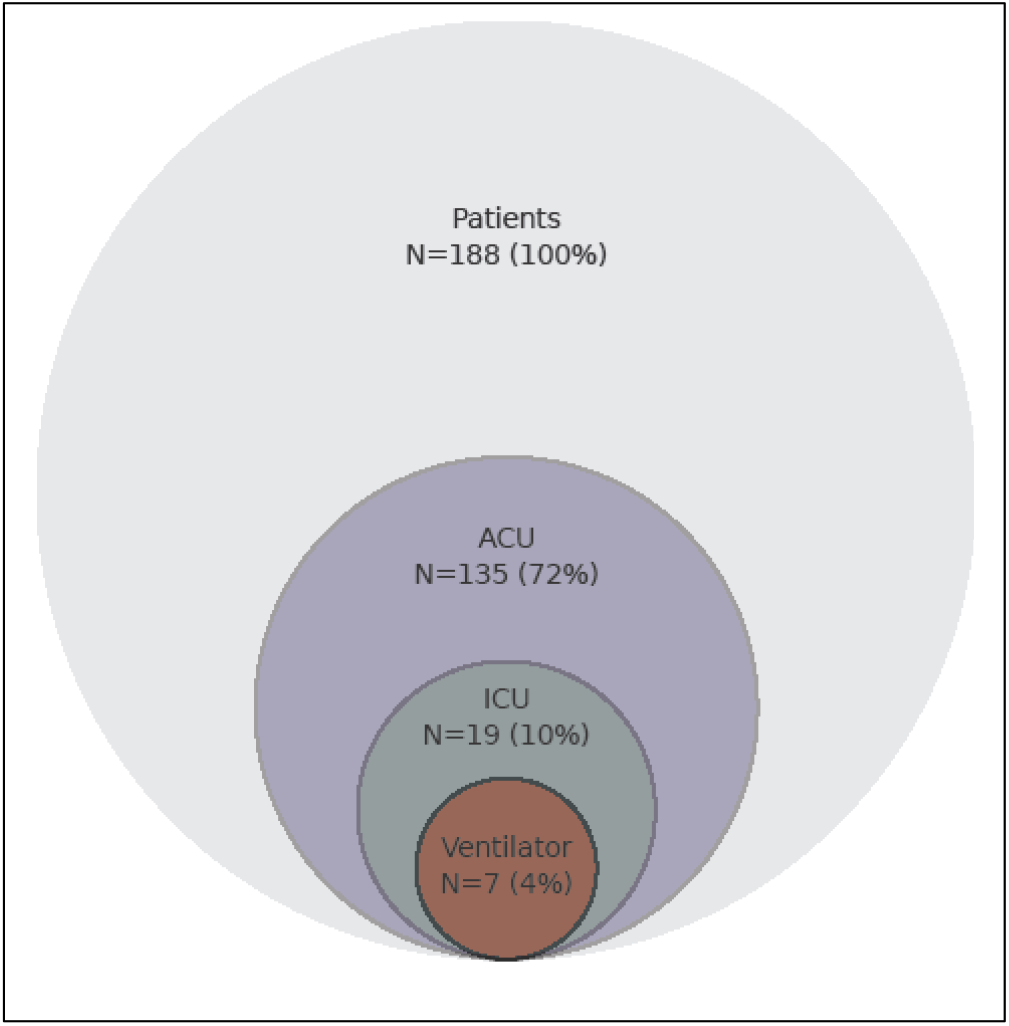
Venn diagram of clinical presentation of COVID-19 patients. The relative frequency of patients requiring different levels of clinical care in the cohort. ACU=Acute care unit. ICU=Intensive care unit.

**Figure 5.**
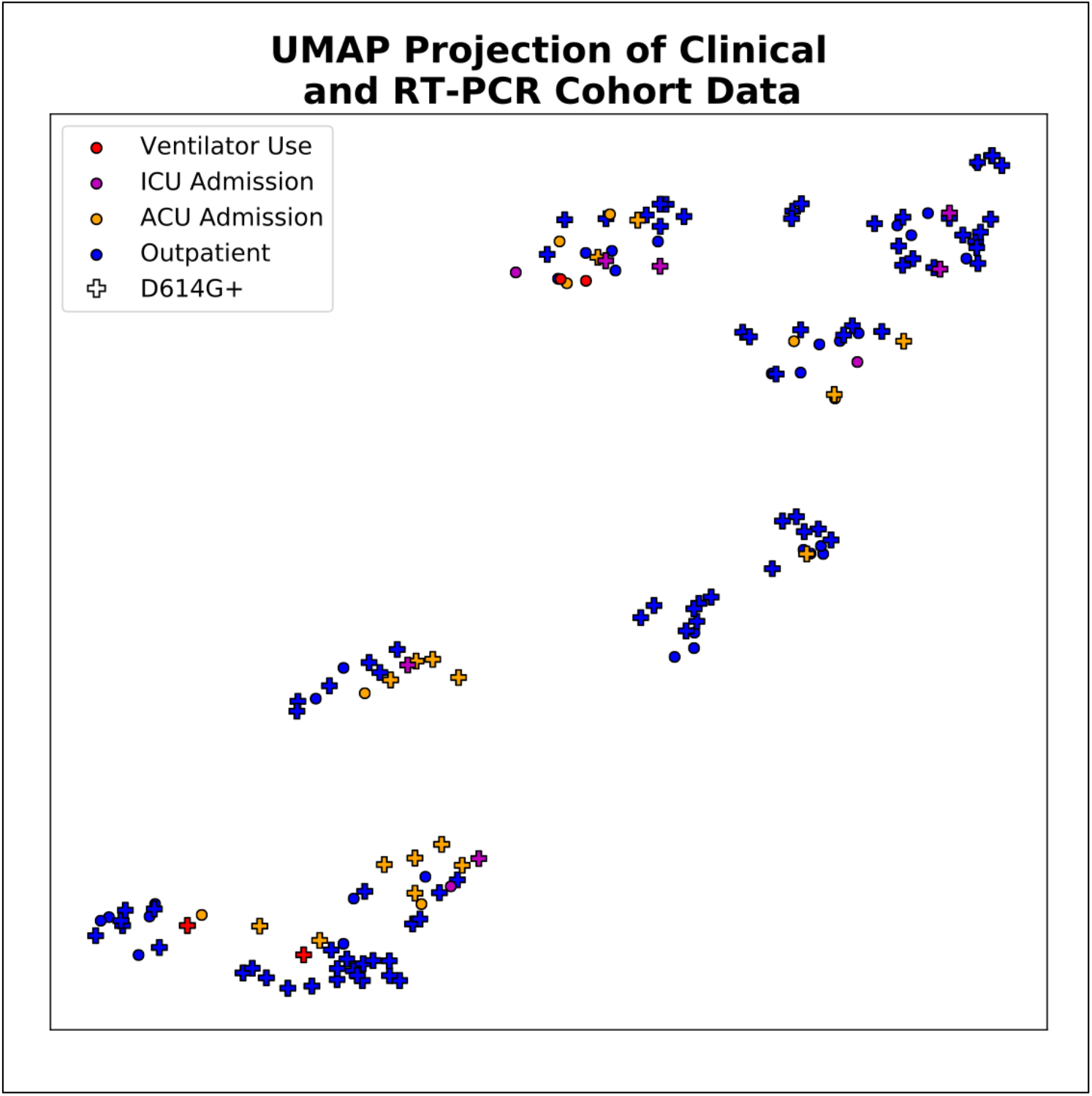
UMAP Projection of EHR and RT-PCR Data. Clinical outcome of a patient is color-coded while D614G status is indicating by a + marker.

We then asked whether SARS-CoV-2 D614G or other genotypes exhibited utility in predicting hospital admission due to COVID-19. We developed a panel of classifiers based on different algorithmic approaches (logistic regression (LR), random forest (RF) and support vector machines (SVM)). For this we divided the sequenced patients into two populations based on admitted versus outpatient. Classifiers were then built based on patient demographics and prior comorbidities alone, demographics plus viral titer in the swab specimen (RT-PCR Ct value), and finally demographics plus viral titer plus D614G genotype. The results for these models based using LR, RF and SVM are detailed in **Figure 6** below. Of note, classification of inpatient versus outpatient was highly accurate based on patient demographics alone using all three approaches, with both the RF and SVM achieving 88% accuracy. Notably, adding viral titer and D614G genotype to the model boosted the accuracy to over 90% for the RF approach, the best performing classifier out of all models tested. From this we conclude that demographic and clinical features have utility for predicting hospital admission in COVID-19 patients, with a modest improvement via the addition of SARS-CoV-2 sample characteristics.

**Figure 6.**
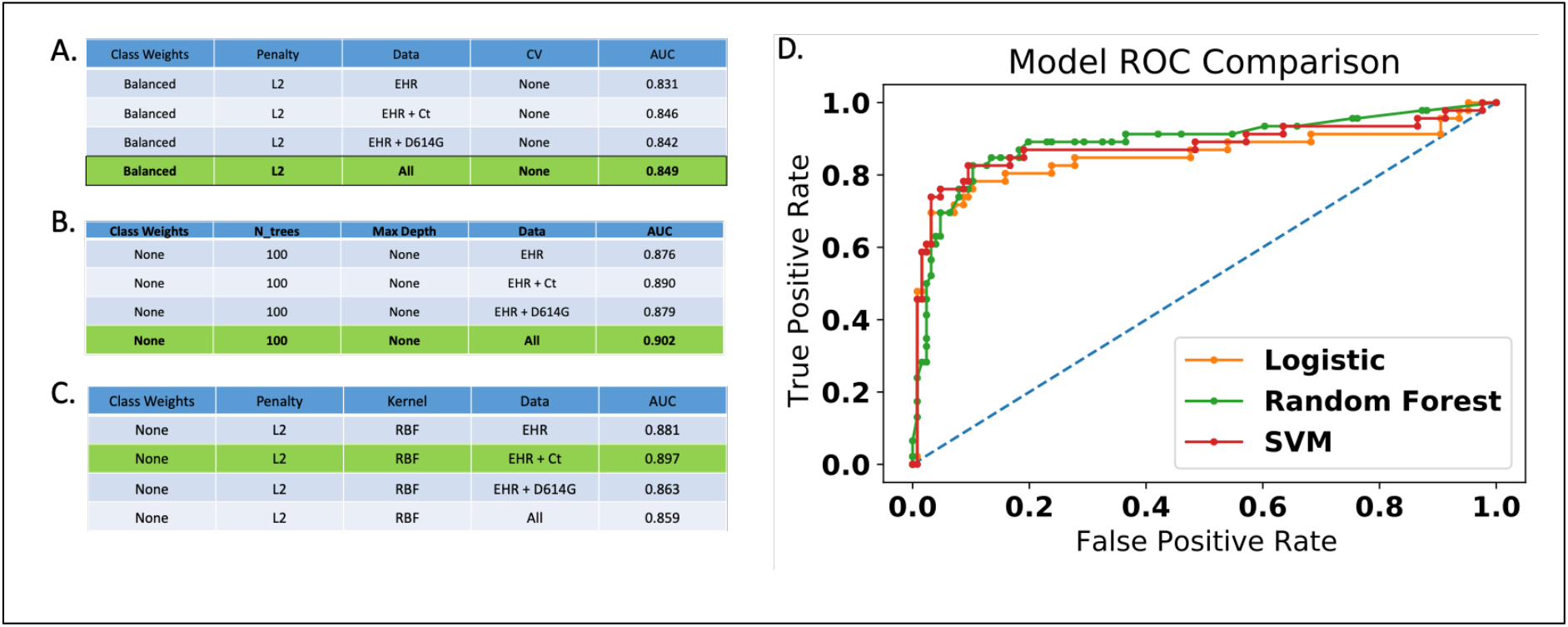
Acute versus non-acute classifier using EHR and genomics. **A)** Acute versus non-acute modeling using a logistic regression approach. **B)** Acute versus non-acute using random forest. **C)** Acute versus non-acute using a support vector machine approach. **D)** Receiver Operator Characteristic (ROC) curves for the three methods.

To further test possible correlations between genotype and patient outcomes, we assessed the predictive value of other prevalent SARS-CoV-2 variants identified in the cohort. For the remaining variants we calculated the Pearson Correlation, variants that were perfectly correlated with each other |*R*_*p*_| = 1 were collapsed into a single statistical test (**Figure 7**). We then tested whether the variant discriminated between outpatients and inpatients using Fisher’s exact test and a logistic regression test (**Table 2**). We note two variants that show some significance across inpatients. The first variant, a T>C mutation at 3312 were significant at p<0.05 in all statistical tests (however we note the small sample size containing this variant). The second variant, an A>G mutation at 20268 shows >95% significance on Fisher’s Exact test with Benjamini-Hochberg, but has poor significance in both logistic regression and permutation tests. Overall, we see modest association with inpatient status across multiple variants; though larger cohort studies are needed to further evaluate this association.

**Table 2.** Viral Variants stratified across Samples, Patients and Outcomes along with Significance Tests for Outpatient vs ACU Admission in Cohort.

| POS | REF | ALT | Patients<br>N=158† | Outpatient<br>N=117† | ACU<br>N= 41 | $p_f^*$ | $p_{fbh}^*$ | $p_{fp}^*$ | $p_{lr}^*$ |
| --- | --- | --- | --- | --- | --- | --- | --- | --- | --- |
| 241 | C | T | 109 (0.69) | 84 (0.72) | 25 (0.61) | 2.40E-01 | 8.07E-01 | 8.57E-01 | 1.98E-02 |
| 1059 | C | T | 40 (0.25) | 29 (0.25) | 11 (0.27) | 8.36E-01 | 9.70E-01 | 3.11E-01 | 1.00 |
| 3312 | T | C | 6 (0.04) | 1 (0.01) | 5 (0.12) | 4.70E-03 | 4.60E-02 | 1.00E-04 | 9.76E-03 |
| 4236 | A | G | 17 (0.11) | 10 (0.09) | 7 (0.17) | 1.47E-01 | 6.72E-01 | 4.20E-02 | 7.00E-01 |
| 11083 | G | T | 38 (0.24) | 23 (0.20) | 15 (0.37) | 3.51E-02 | 2.75E-01 | 1.11E-02 | 1.00 |
| 11563 | C | T | 22 (0.14) | 14 (0.12) | 8 (0.20) | 2.93E-01 | 8.24E-01 | 7.42E-02 | 1.00 |
| 14805 | C | T | 37 (0.23) | 23 (0.20) | 14 (0.34) | 8.51E-02 | 5.09E-01 | 2.19E-02 | 1.00 |
| 18086 | C | T | 11 (0.07) | 9 (0.08) | 2 (0.05) | 7.30E-01 | 9.70E-01 | 5.79E-01 | 9.55E-01 |
| 18877 | C | T | 27 (0.17) | 19 (0.16) | 8 (0.20) | 6.35E-01 | 9.70E-01 | 2.33E-01 | 1.00 |
| 20268 | A | G | 18 (0.11) | 18 (0.15) | 0 (0.00) | 3.95E-03 | 4.26E-02 | 9.97E-01 | 9.99E-01 |
| 25563 | G | T | 67 (0.42) | 48 (0.41) | 19 (0.46) | 5.85E-01 | 9.70E-01 | 2.19E-01 | 1.00 |

† Values are numbers (percentages).
\* $p_f$ : Fisher's Exact Test; $p_{fbh}$ : $p_f$ with Benjamini-Hochberg correction;
$p_{fp}$ : $p_f$ with permutation adjustment; $p_{lr}$ : Logistic Regression Test

**Figure 7.**
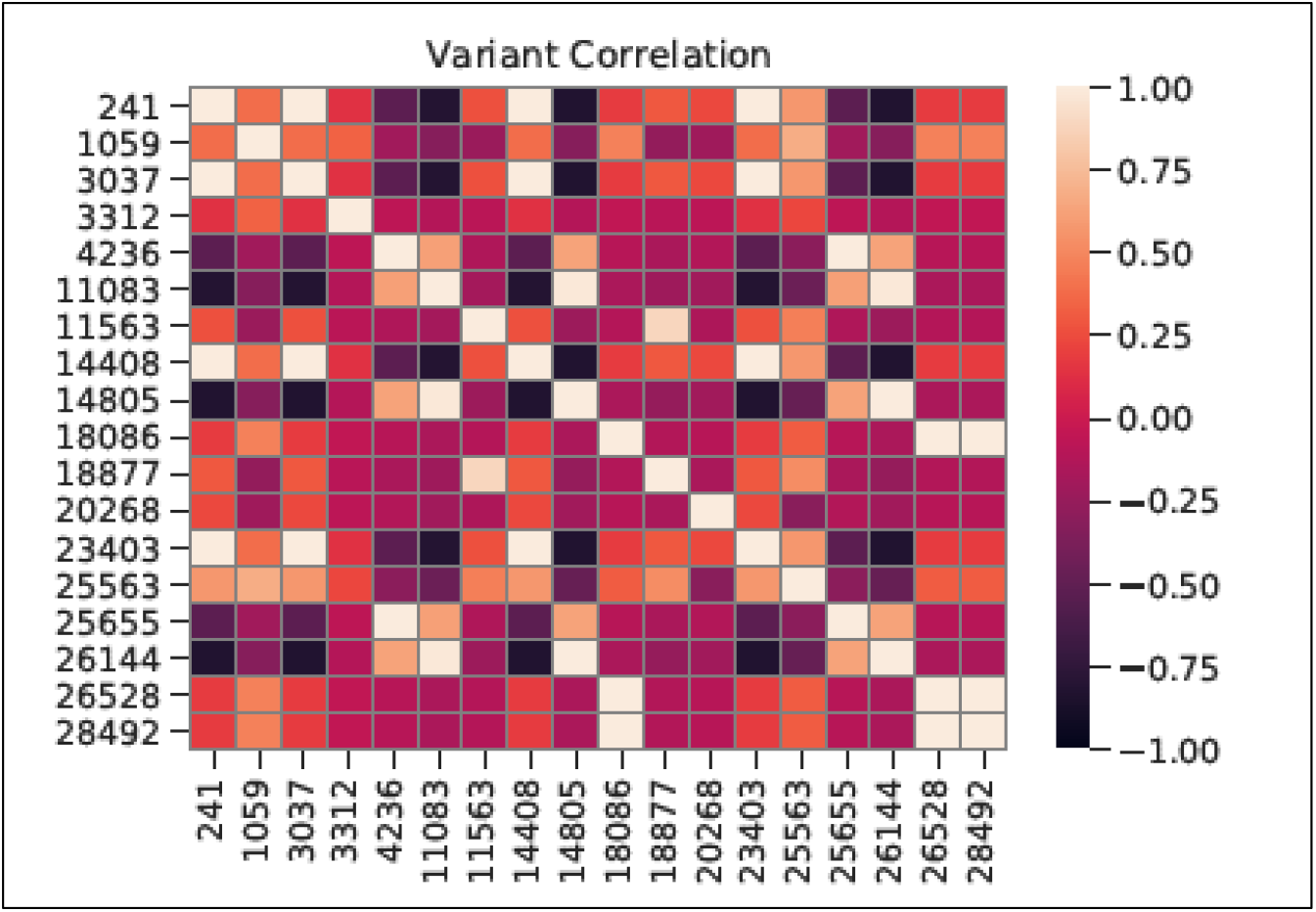
Heatmap Showing the Pearson Correlation among variants that appear with a Frequency > 11 in Viral Genomic Samples.

## DISCUSSION

This study represents a genomic catalog of SARS-CoV-2 introduction and spread throughout Oregon with an analysis of specimens dating to just days after the initial known first case through May 2020. Unlike the earlier outbreak in neighboring Washington State starting in late January 2020 (Bedford et al., 2020) that was hallmarked by high homogeneity, early cases in Oregon exhibit striking diversity indicative of multiple introduction events into the state, similar to patterns others have elucidated for neighboring northern California (Deng et al., 2020). Given the widespread nature of SARS-CoV-2 outbreaks across the United States in March, with some states such as New York reporting tens of thousands of cases by end of month, it is perhaps unsurprising that early cases of SARS-CoV-2 in Oregon represent a diverse set of most major clades. The change over time in OR to a prevalence of D614G-positive infections also mirrors a trend observed worldwide as well as biological data supporting increased infectivity due to this alteration (Grubaugh et al., 2020; Korber et al., 2020; Zhang et al., 2020); it is somewhat remarkable that this shift is dramatically observable in smaller sample numbers in a localized area.

Overall, we show that hospital admission in COVID-19 cases is predictable (>90% predictive model classification accuracy, Random Forest approach) using prior clinical information and viral specimen information. Of these data types, patient demographics such as age as well as clinical priors and comorbidities were clearly the main contributors, as was expected. However, it is important to note that adding in SARS-CoV-2 genomic information did reproducibly improve prediction accuracy tested across a significant number of bootstrap iterations. While D614G has not yet been shown to be associated with differences in clinical symptoms aside from nasal viral titer in other studies, it is of note that in our study, eleven out of the twelve patients that died due to COVID-19-releated complications were infected with D614G-negative SARS-CoV-2 virus. While our models accounted for differences in patient age, we note the small sample size and also note that it is difficult to decouple additional confounders such as changes in treatment strategies and emergence of new therapies that have rapidly occurred over time during the COVID-19 crisis. It will be important to test these associations across larger cohorts of genomic and associated clinical data.

It is also important to note that the majority of patient samples assessed in this study had one or more alterations that made them either completely unique or shared with only a few additional samples in the cohort. While SARS-CoV-2 sequence data are not yet widely utilized for outbreak tracing, our study clearly shows remarkable utility for these purposes. Given the sheer number of circulating rare alterations, there is perhaps no other methodology that can conclusively confirm or rule out a unique person-to-person transmission event. Given the small number of sequencing reads required to sequence a SARS-CoV-2 genome and the high-throughput capacity of modern next-generation sequencers, this strategy remains a relatively inexpensive and scalable tool that should be under consideration for all viral outbreak tracing programs worldwide.

## MATERIALS AND METHODS

### Patients

All studies were conducted on discarded specimens and associated deidentified clinical information under IRB approval (study protocol 2020000127) with approved waiver of consent.

### Specimens

Discarded nasopharyngeal swab specimens previously assessed as positive for SARS-CoV-2 by RT-PCR using either a Cobas SARS-CoV-2 RT-PCR assay on a Cobas 6800 system (Roche Diagnostics) or a BioGX SARS-CoV-2 assay on a BD MAX system (Becton Dickinson) were selected for sequencing. Specimens were suspended in 3 mL of Viral Transport Medium (VTM) (Hanks’ Balanced Salt Solution [HBSS] supplemented with 2% heat-inactivated fetal bovine serum [FBS], 100 µg/mL Gentamicin, and 0.5 µg/mL Amphotericin B), phosphate-buffered saline (PBS) or Copan Universal Transport Medium (UTM). Specimens were collected over a period of March to May 2020.

### Nucleic Acid Purification

A mixture of total nucleic acids (NA) was isolated from 200 µL of swab-inoculated media using the QIAsymphony DSP Virus/Pathogen (Mini or Midi) kits and a QIAsymphony SP instrument (Qiagen). NA purification took place following the addition of carrier (poly-A) RNA and an internal control RNA, according to the manufacturer’s instructions. Purified NA were eluted in 85 µL of Elution Buffer AVE (Qiagen; RNase-free water with 0.04% NaN_3_).

### Sequencing Library Preparation

Sequencing libraries were prepared using Illuminia TruSeq RNA Library Prep for Enrichment kits (Illumina), and SuperScript II Reverse Transcriptase (ThermoFisher Scientific). Given the challenge of quantifying SARS-CoV-2 RNA within the purified NA, 8.5 µL of QIAsymphony eluate was used as starting material for library preparation. When samples possessed a SARS-CoV-2 threshold cycle (C_t_) value of lower than 10.2, a 10-fold dilution of purified NA was performed prior to library preparation.

### Hybrid Capture

Indexed cDNA libraries were pooled in groups of eight; each pool contained a total of 1.5 µg. Such pools were constituted of samples having similar SARS-CoV-2 C_t_ values (cobas 6800, Target 1, Roche). SARS-CoV-2 cDNAs were enriched using a hybrid-capture oligonucleotide panel covering the SARS-CoV-2 genome (GenBank Accession MN908947.3; 29,903 base pairs [bp]; Twist Bioscience) and a commercial hybrid-capture kit (Twist Bioscience). This approach yielded adequate (>60x average per base) coverage for clinical specimens with an initial RT-PCR cycle threshold of 30 or below (**Figure S1**).

### DNA Sequencing

SARS-CoV-2-enriched libraries were sequenced on a NovaSeq 6000 or iSeq 100 instrument (Illumina) using paired-end reads of 150 bp each.

### Sequencing Data Analysis

Raw sequencing reads were mapped against the reference sequence MN908947.3 using BWA-MEM v0.7.12-r1039 (Li and Durbin, 2009). Mapping statistics were calculated via samtools flagstat (samtools v1.10) (Li et al., 2009). Coverage was assessed by generating a coverage plot for each sample illustrating percent coverage across the genome via deepTools plotCoverage v3.4.3 (Ramirez et al., 2014). Samples with 60x or less coverage at the 90% percentile were excluded from downstream analysis. Average coverage for the 179 samples passing filter was approximately 3000x. Variant calling was performed using Freebayes v1.3.2-40-gcce27fc with standard filters and ploidy of 1. Variant calls were further filtered via bcftools vcffilter (bcftools v1.10.2-61) with parameters requiring a quality score > 3000, minimum read depth > 100, and minor allele frequency >= 0.5. Remaining variants passing filter were annotated via SnpEff v4.3 (Cingolani et al., 2012). The resulting variant calling file was used in conjunction with reference genome MN908947.3 to construct a consensus genome for each sample via bcftools consensus. Consensus sequences were manually assessed for variant call accuracy using Geneious Prime v2020.1.2 to confirm accuracy before submission to GISAID. SARS-CoV-2 genotype diversity was assessed by extracting variant calls from annotated VCFs using a custom Python script and generating plots using custom R scripts.

### Variance Significance

Variants from Variant Call Files (vcfs) are mapped into an *N*_*s*_ *× M* indicator matrix is where *N*_*s*_ is the number of samples while *M* is the number of unique variants in the sample set. Initially, *N*_*s*_ = 181 and *M* = 233. We filter out all variants that do not appear in at least 11 samples. On the remaining variants we calculate the Pearson Correlation matrix and find any groups of variants that are perfected correlated or anticorrelated. From these groups we filter out all but the first variant. Significance (*p* values) is first calculated using Fisher’s Exact Test for two-tailed distributions. To correct for multiple-hypothesis tests we adjust the *p* values using the Benjamini-Hochberg correction with a family-wise error rate of 0.1. For further validation, we apply a permutation correction (with number of permutations= 20000) along with Fisher’s Exact Test to correct the *p* value. Finally, we calculate a final set of *p* values using a logistic regression model.

### Clinical Prediction and Analysis

Patient’s age and Ct values have been normalized to be [0,1] by divind by either the max Ct value or dividing by 100 for the patient’s age. Current and former smoking status was merged into a single categorical variable. Finally, we removed any categorical variable whose total sample size < 5. For the UMAP projection we set *N*_*neighbors*_ = 15, *min_distance* = 0.1, and used the Euclidean distance metric. All predictive models (LR, SVM, RF) where trained using the *Scikit-Image* library.

## DATA SHARING

All SARS-CoV-2 sequences generated for this manuscript are publicly available through GISAID.org (under originating laboratory “Providence St. Joseph Health Molecular Genomics Laboratory” or by author filter for “Dowdell *et al*.”). Deidentified clinical data are available upon request.

**Figure S1.**
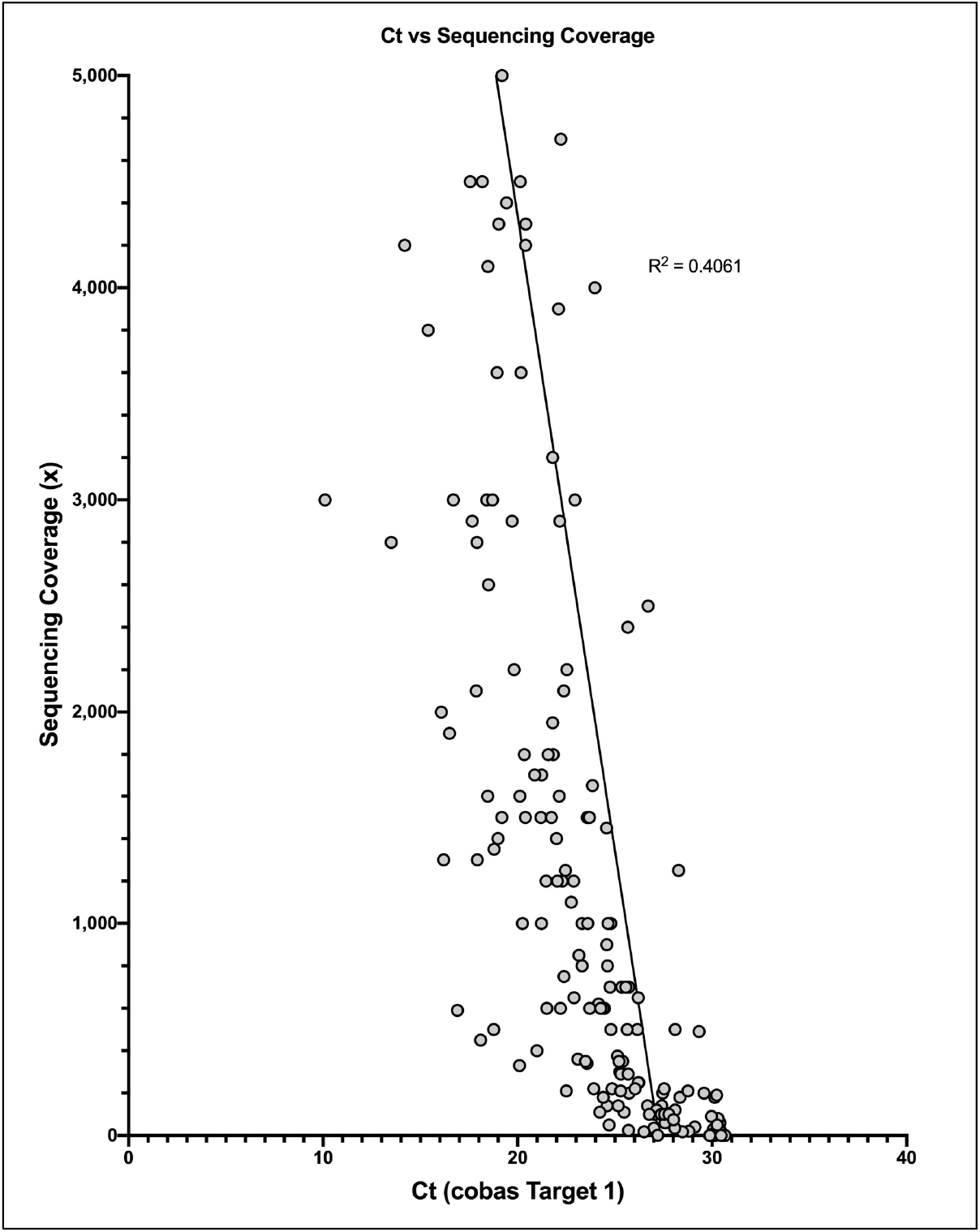
Correlation between RT-PCR cycle threshold and sequencing genomic coverage.

## Data Availability

All SARS-CoV-2 sequences generated for this manuscript are publicly available through GISAID.org (under originating laboratory Providence St. Joseph Health Molecular Genomics Laboratory or by author filter for Dowdell et al.). Deidentified clinical data are available upon request.

http://www.gisaid.org

